# Predicting cognitive impairment using novel functional features of spatial proximity and circularity in the digital clock drawing test

**DOI:** 10.64898/2026.03.14.26348336

**Authors:** Adlin Pinheiro, Cody Karjadi, Yorghos Tripodis, Vijaya B. Kolachalama, Kathryn Lunetta, Serkalem Demissie, Chunyu Liu, Rhoda Au, Shariq Mohammed

## Abstract

The digital clock drawing test (dCDT) is a cognitive screening tool employing a digital pen. While many studies rely on summary statistics of dCDT features to predict cognitive outcomes, these approaches often involve subjective decisions such as feature selection and imputation. In this study, we introduce novel dCDT features, expressed as mathematical functions, and compare them to commonly used summary features. We included dCDTs from 3,415 participants from the Framingham Heart Study. Random forest models with five-fold cross-validation were trained to distinguish participants with mild cognitive impairment or dementia from cognitively intact participants. When combined with established time-based features, functional features related to spatial proximity and circularity demonstrated predictive power comparable to commonly used summary features. Our findings highlight the potential of integrating functional features to detect subtle motions and behaviors in digital cognitive assessments, offering new tools that may enhance diagnostic accuracy and support early detection strategies.

## 1 Introduction

The Clock Drawing Test (CDT) is a widely used screening tool within standard neuropsychological assessments, evaluating cognitive domains such as memory, visuo-spatial abilities, and executive control functioning [1, 2]. During the test, individuals are asked to draw a clock from memory, positioning the hands to depict ten past eleven (command condition). Additionally, in some settings, either immediately after or after a short time delay, participants are asked to replicate an image of a clock provided to them (copy condition). The CDT is relatively straightforward, cost-effective, and can be administered in diverse settings. Moreover, it has been shown to effectively detect moderate to severe dementia [2]. However, its performance varies when identifying mild dementia or mild cognitive impairment (MCI) [3–6] and distinguishing between different dementia subtypes [7].

Recent advances in digital pen technology have enabled a digital version of the CDT (dCDT) that reduces inter-rater variability and the subjectivity inherent in manual scoring [8]. The digital Clock Drawing Test (dCDT) records pen position, pressure, and time at high frequencies–every 13 milliseconds–with spatial accuracy of approximately ±0.002 inches [9]. Consequently, dCDT detects subtle pen movements and behaviors that were previously unobservable with traditional paper-and-pen–based tests (e.g., time interval between strokes, total duration the pen is in contact with the paper). Around 100 latencies and graphomotor features have been extracted from the dCDT across studies [10]. The features derived from the dCDT have demonstrated improved diagnostic accuracy in distinguishing cognitive groups compared to traditional CDT scoring systems [11]. Moreover, multiple studies report that dCDT-based measures distinguish cognitively intact individuals from those with cognitive impairment [8, 12–17].

A variety of features derived from dCDT have been employed in statistical and machine learning models to classify cognitive outcomes [8, 12–14]. One such approach, DCTclock, uses measurements derived from the final drawing and the drawing process to form composite scores representing cognitive function [8]. This approach has been shown to distinguish between cognitively intact and impaired individuals [8, 16] and has been validated against positron emission tomography (PET) biomarkers associated with Alzheimer’s Disease (AD) [16].

However, selecting and aggregating large sets of hand-engineered features introduces analyst subjectivity, necessitates careful handling of feature correlations, and requires imputation of missing values. Collapsing rich trajectories into a limited set of summaries can also obscure informative structure in the drawing process. For instance, clock-face summaries usually include symmetry, area, and clock-face overshoot distance/angle [8, 14, 18], and pressure is commonly summarized by the mean, standard deviation, or a pressure-velocity ratio [12, 14]. Such summarization underscores the need for representations that retain the temporal and geometric detail present in the raw pen traces while remaining interpretable and tractable for modeling.

### 1.1 Novel Functional Features

To address these limitations, we developed novel features represented as mathematical functions. Specifically, we construct three features: a nearest-neighbor *G*-function to summarize spatial proximity of observed pen points, a radius function to quantify clock-face circularity, and a pressure density function to characterize the distribution of the pressure applied. These features are designed to reveal structural nuances of the dCDT by identi-fying subtle variations and patterns in the drawing process, while remaining interpretable. Moreover, these novel features can be computed even when the test/drawing is incomplete (e.g., missing digits or hands, or a partially drawn clock-face), reducing reliance on ad-hoc imputation.

### Motivation for the choice of features

The three functions we consider target complementary aspects of the drawing: where and how long participants dwell spatially (*G*-function), how the clock-face is shaped (radius), and how force is applied (pressure density).

- Studies have shown evidence that individuals with dementia exhibit longer completion times [12, 14, 18], reduced drawing speed [12], and smaller clock-face areas [17, 18]. *G*-function serves as a single, interpretable functional summary of spatial behavior rather than treating these correlates separately.
- Clock-face shape and symmetry have been previously used in classification models to predict cognitive impairment [8, 14, 17] and has shown promise. Therefore, we chose the radius function to capture deviations from circularity in a continuous, comprehensive and interpretable way.
- Evidence on pen pressure in the dCDT is mixed. One study found that mean, standard deviation, and the pressure-velocity ratio were highly informative, with dementia cases showing lower mean pressure in both conditions and lower variability in the command condition [14]. Similar reductions have been observed in other non-clock digital drawings from AD patients [19]. In contrast, another study found no significant differences in mean pressure and pressure-velocity ratio across healthy controls, amnestic MCI, and mild AD [12].

We reduce each function to a small number of scores via functional principal component analysis[20] and evaluate their predictive performance against established summary-feature models in a large, community-based cohort. In a large cohort of community-dwelling individuals, we demonstrate that features capturing spatial proximity (*G*-function) and circularity (radius) achieve performance comparable to models built from traditional summary features, whereas pressure density contributes limited incremental value. These advancements offer a more nuanced understanding of subtle cognitive behaviors, paving the way for enhanced diagnostic precision and early intervention in cognitive impairment.

An overview of the end-to-end workflow is shown in Fig. 1. In Section 2, we describe the cohort, outcomes, and pre-processing, followed by details on the functional feature construction, dimension reduction, and the cross-validated prediction modeling framework. In Section 3, we report sample characteristics, functional pattern summaries, and classification results, and include sensitivity analyses and variable-importance findings. In Section 4, we interpret the results, discuss main finding and note limitations.

**Figure 1:**
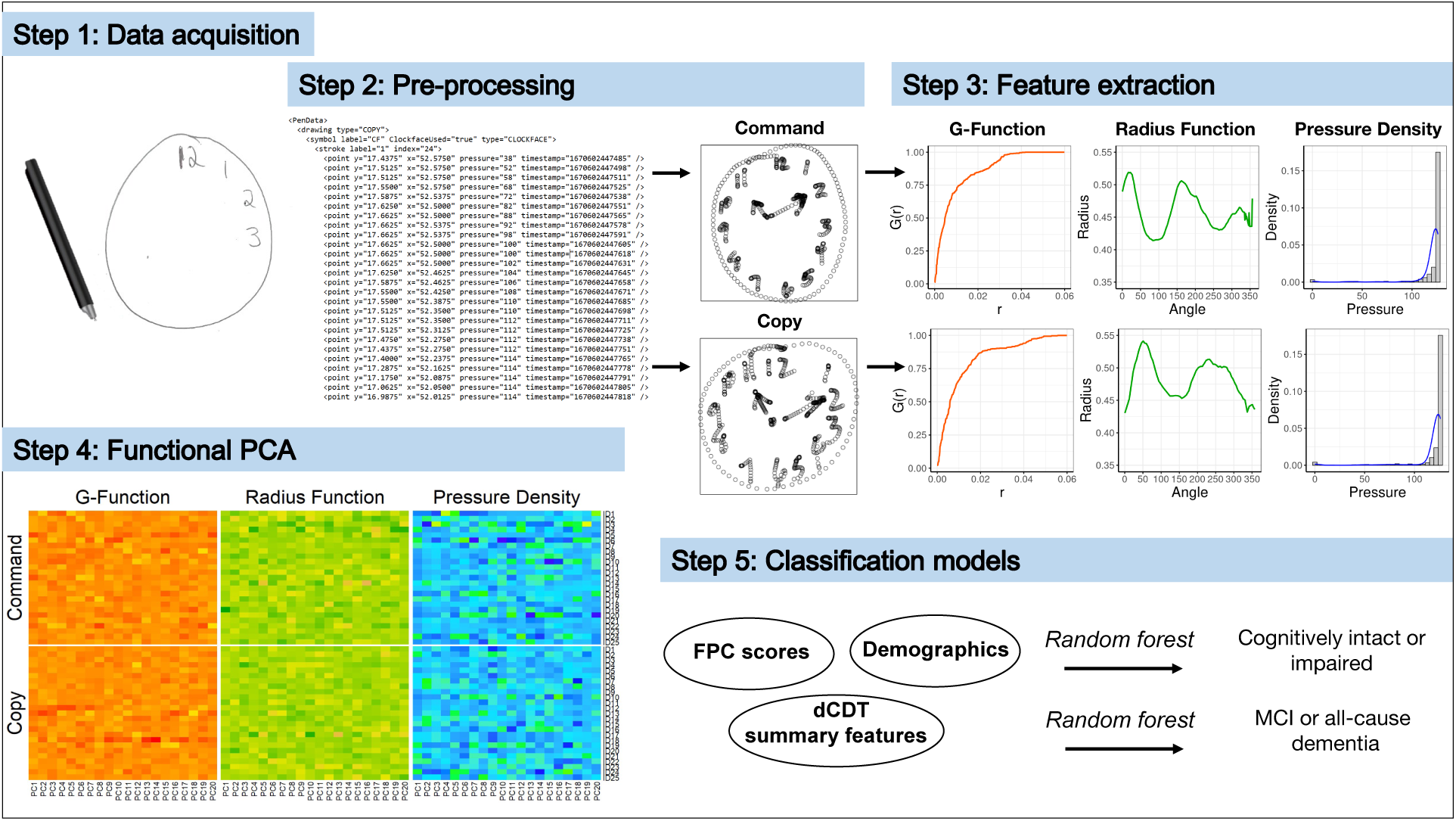
Overview of the analysis pipeline. *Step 1 — Data acquisition:* Digital pen records (*x, y*), pressure, and timestamps for command and copy dCDTs. *Step 2 — Processing:* Basic scaling/translation to a common coordinate system. *Step 3 — Functional feature extraction: G*-function (spatial proximity), radius function (circularity), and pressure density. *Step 4 — Functional PCA:* Dimension reduction to a small set of functional principal component (FPC) scores. *Step 5 — Classification:* Random-forest models evaluated by AUC, sensitivity, and specificity.

## 2 Methods

### 2.1 Data and Pre-processing

#### Participants

The Framingham Heart Study is a prospective, multigenerational, community-based cohort study established in Framingham, Massachusetts in 1948 with the recruitment of the Original cohort [21]. Subsequently, recruitment began in 1971 for the Offspring cohort, consisting of the children and their spouses of the Offspring cohort. In 1995, the Omni 1 cohort was recruited, consisting primarily of minorities to reflect the changing demographics of Framingham, MA. Children of the Offspring cohort were recruited for the Third Generation cohort in 2002 as well as a second minority cohort, Omni 2.

The CDT is administered as part of standard neuropsychological testing within the FHS. In 2011, the FHS transitioned from the traditional paper and pen method to digital pen technology for the CDT. Participants typically complete both the command and copy conditions of the test. The accompanying software automatically classifies each stroke into clock components (e.g. clock-face, minute/hour hand, digit, etc.) with a research assistant manually correcting classification errors as needed.

Through the end of 2021, there are 4,959 dCDTs from 3,701 participants. For the present study, we considered only the most recent test from participants who completed either a command or copy clock. Participants were excluded if they had a test with pen malfunctions, were non-native English speakers, or had missing clinical characteristics, poor vision, recent head trauma, or history of stroke. The final sample comprised 3,415 participants.

The Institutional Review Board of Boston University Medical Center approved the study protocol and informed consent was obtained from all participants.

#### Outcome Measures

Certain participants were flagged for an adjudicated review process conducted by a dementia review panel, which included a neurologist and a neuropsychologist. Diagnoses followed DSM-IV criteria [22] classifying participants as cognitively intact, MCI, or dementia (including AD) [23]. The cognitively intact group is defined as those participants who were either not selected for adjudicated review or were deemed cognitively intact upon review. The diagnosis of MCI or dementia was made either prior to the date of the dCDT or within 180 days after. The primary outcome contrasted cognitively impaired (MCI or dementia) versus cognitively intact; the secondary outcome contrasted all-cause dementia versus MCI.

#### dCDT Pre-processing

Raw dCDT files contain high-resolution pen movement data captured every 13 milliseconds. Each recorded point of the digital pen contains the *x* and *y* coordinates, pen pressure, timestamp, stroke label, component label, and test condition. To place drawings across participants on a common geometric scale, we linearly scaled and translated the coordinates to the unit square, preserving aspect ratio and maintaining the relative Euclidean distances between points. This subject-specific coordinate scaling factor (reflecting overall drawing size)was included as a covariate in downstream prediction models. Figure 1 provides an overview of the pre-processing and analysis pipeline.

### 2.2 dCDT Functional Features

Recent studies have defined and used various features of the dCDT to predict cognitive outcomes. These collections of hand-engineered *summary features*, can be broadly categorized into measures that capture drawing efficiency (e.g., total time, number of strokes), information processing (e.g., think time, latency), spatial reasoning (e.g., missing digits/hand, clock-face area), and simple and complex motor features (e.g., ink time, speed) [8, 16]. Table S1 summarizes the set of summary features we implemented from the literature. We compare predictive performance of the summary features to our novel, interpretable functional features that retain temporal and geometric detail and are computable even when drawings are incomplete.

#### Nearest-Neighbor *G*-function (Spatial Proximity)

*G*-function quantifies the spatial distribution of points within a closed area. It is the empirical cumulative distribution function of nearest-neighbor distances between points, giving the probability that a randomly chosen point has at least one other point within radius *r*. The *G*-function is computed as 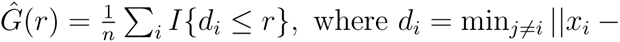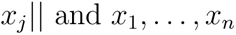 are the *n* points drawn in the test. It provides insights into the level of clustering or dispersion of points within an area. For example, a rapidly rising *G*-function indicates a tightly clustered spatial point pattern compared to a moderately increasing one that indicates dispersion of points.

In the dCDT, points are sampled at a constant interval; thus the spatial clustering captured by *G*(*r*) reflects a joint signature of *where* participants draw and *how long* they spend in a local region. Specifically, the *G*-function reflects the complex interplay between the time taken to complete the test, speed, and the spatial layout of the points. For the dCDT, a more rapidly rising *G*(*r*) indicates tighter clustering (e.g., slower or more hesitant movements within a smaller area), whereas a flatter curve indicates more dispersed points (e.g., faster pen movements).

We compute *G*(*r*) from all recorded points, independent of stroke/component labels, which reduces dependence on stroke-classification accuracy. It was evaluated at a fixed, dense grid of 500 equally-spaced *r* values, from 0 to *r_max_*. Here *r_max_* is the largest radius value considered when analyzing the spatial distribution of points across all tests. The R package spatstat was used to compute the *G*-functions.

#### Radius Function (Circularity)

To quantify the circularity of the clock-face comprehensively, we define radius function as the distance from each point on the clock-face to its center as a function of the angle (formed by a reference line that starts at the 12 o’clock position–vertical from the center– and rotates clockwise from 0 to 360 degrees). The radius function includes only those points from strokes classified by the software as originating from the clock-face.

A clock-face that is a perfect circle yields a flat radius function. Deviations from a flat function encode asymmetry, warping, and local imprecision. This functional view generalizes discrete symmetry or area summaries into a shape signal that is still clinically interpretable.

Because overlapping strokes can cause self-intersections, we first form the convex hull of clock-face points to obtain a non-self-intersecting boundary; we then augment hull vertices with additional face points at unique angles and linearly interpolate to obtain a radius value at each integer degree. Convex hulls were also used to estimate clock-faces for nine participants with partially drawn or missing clock-faces to provide an approximate boundary reflecting the intended clock shape. Further details about the data processing and computation of the radius function are provided in the Supplementary Materials.

#### Pressure Density Function

To obtain a more comprehensive understanding of pressure’s impact on cognition, we model the full distribution of pressure values via a nonparametric density function rather than relying on limited summary measures such as mean and standard deviation. The digital pen records the pressure applied to the paper at each point, with values ranging from 0 (no pressure) to 126 (maximum pressure). The probability density function of these pressure values captures the entire distribution during the test, providing detailed insights into the heterogeneity of pressure applied.

For each test, the density function was estimated non-parametrically using a Gaussian kernel for smoothing. Similar to the *G*-function, the pressure density function does not rely on stroke labeling. It was evaluated on a dense grid of 500 equally-spaced values ranging from 0 to 126.

### 2.3 Functional Data Analysis

Functional data provide information about curves and surfaces defined over a specific domain of interest. Each observation/sample in functional data is viewed as discrete realizations of an unknown, smooth random function in a stochastic process, observed with or without error. For a sample of *N* functions, the function corresponding to the participant *i* ∈ {1*, . . ., N* }, denoted as *x_i_*(*t*), is observed at 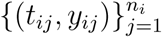 with *t_ij_* ∈ [*T*_1_*, T*_2_] and *y_ij_* ∈ R, allowing *n_i_* and sampling locations to vary across *i*. When data are observed with error, the observed data can be represented as *y_ij_* = *x_i_*(*t_ij_*) + *ɛ_ij_*, where *ɛ_ij_* represents the error observed at *t_ij_* and is assumed to be independent and identically distributed with *E*(*ɛ_ij_*) = 0 and Var(*ɛ_ij_*) = *σ*^2^. Our goal is to obtain low-dimensional, interpretable summaries of *x_i_*(·) to use as predictors.

The fundamental idea in functional data analysis (FDA) is that the function corresponding to participant *i* can be expressed as a linear combination of *K* basis functions.[24] Let *ϕ*(*t*) = (*ϕ*_1_(*t*)*, . . ., ϕ_K_*(*t*))^⊤^ denote a chosen basis; then

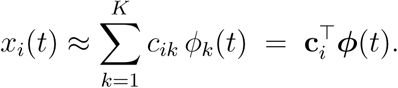

When data are observed without error, coefficients can be obtained by interpolation. However, when data are observed with error, the degree of smoothing is controlled by the number of basis functions and the corresponding coefficients are determined by least squares estimation [24].

#### Functional Principal Components Analysis (FPCA)

Because the proposed features are functions (infinite-dimensional objects), they cannot be directly used as predictors/features in standard statistical or machine learning models. Hence we perform dimension reduction so they can be integrated used as predictors. A common tool for this purpose in FDA is the FPCA. Analogous to multivariate principal component analysis, FPCA aims to preserve as much variability in the original functions as possible while exploring the primary modes of variation in the sample of functions to reduce data dimensionality.

In multivariate PCA, for an *n* × *p* data matrix *X* with covariance matrix *V*, we identify weight vectors *ξ* that maximize the variance of the projections *V ar*(*ξ*^⊤^*X*) = *ξ^T^ V ξ*. This leads to the eigen-decomposition *V ξ* = *λξ*. FPCA extends this logic to continuous trajectories. Let *µ*(*t*) = E{*x_i_*(*t*)} and *G*(*s, t*) = Cov{*x_i_*(*s*)*, x_i_*(*t*)}. FPCA seeks weight functions that maximize the variance of the scores

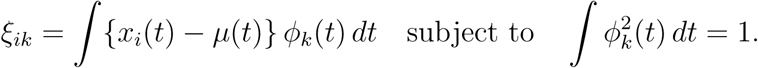

The optimal weight functions (eigenfunctions) *ϕ_k_*(·) and their associated variances (eigen- values) *λ_k_*are found by solving the functional analogue of the eigen-equation

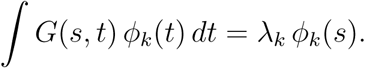

Under the Karhunen–Lòeve representation [24], we can then represent each *x_i_*(*t*) as

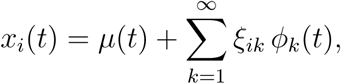

where he first *K* FPC scores, *ξ_i_*_1_*, . . ., ξ_iK_*, capture most variability and serve as low-dimensional, interpretable features.

Principal components analysis through conditional expectation (PACE) is an alternative approach to estimate principal components scores of functional data, based on certain additional assumptions [20]. This method is specifically designed to handle sparse and irregularly sampled data, and it has been demonstrated to enhance traditional FPCA approaches for densely sampled functional data. Moreover, the individual curves do not require pre-processing or smoothing prior to analysis. The PACE framework has similar setup, where the observed data are independent realizations of a smooth random function, with an unknown underlying mean function *µ*(*t*) and covariance function *G*(*s, t*). The estimates for the mean, *µ̂*(*t*), and covariance functions, *Ĝ*(*s, t*), are derived by smoothing the covariance function from the observed data using local linear and quadratic smoothers, respectively. The eigenfunctions and eigenvalues of the subsequent eigendecomposition are estimated by discretizing the smoothed covariance function. The analytical form of the FPC scores is provided in the Supplementary Materials. Additional assumptions on the distribution of error terms and FPC scores facilitate the management of sparse data via the PACE method [20]. In the analysis presented in this paper, we employ the FPC scores calculated by the PACE algorithm.

### 2.4 Prediction Modeling Framework

Baseline clinical characteristics, including age, sex, education, FHS cohort, and the presence of the APOE4 allele were obtained for the entire sample; we summarized the distribution of these variables by cognitive status. Our primary classification task involves distinguishing between cognitively impaired and cognitively intact participants. The cognitively impaired group included individuals with either MCI or dementia, reflecting the use of the CDT as a screening tool for cognitive impairment rather than a diagnostic tool. A secondary classification task was conducted to classify participants with dementia from those with MCI.

### Model families and setup

We trained random forest classifiers using stratified five-fold cross-validation to preserve class proportions within each fold. For both the primary and secondary classification tasks, we evaluated the following pre-specified model families.

- *Demographics only*: age, sex, education, FHS cohort, and APOE-*ε*4 status.
- *Demographics + summary features*: demographics plus literature-based dCDT summary measures.
- *Demographics + time + single functional feature*: demographics; time-based measures (total completion time, total ink time, total think time); and *one* functional feature (FPC scores from the *G*-function, radius function, or pressure-density function). Whenever a functional feature was included, we also added the image scale factor.
- *Demographics + time + all functional features*: demographics; time-based measures; FPC scores from *all three* functional features (*G*, radius, pressure density); plus the image scale factor.

Each model family was fit separately for command-only, copy-only, and combined command+copy inputs. Note that the *Demographics + summary features* model is used to benchmark against existing practice. The summary features are a literature-aggregated set collated from prior dCDT studies (Table S1) and analyzed under the same modeling pipeline. This does not reproduce any single published model; instead, the collated set is evaluated with the same leakage-controlled, stratified five-fold cross-validation used for all model families. As such, it provides a strong and fair comparator for summary-feature approaches in this dataset.

### Functional features and leakage control

The functional features were incorporated into the models by first conducting FPCA using the PACE algorithm on the training set within each fold of the cross-validation process (to avoid information leakage). The functions in the validation set were subsequently projected onto the training-derived eigenfunctions to obtain their FPC scores. he number of principal components *K* for each functional feature was chosen within the training fold to explain at least 99% of total variance. R package fdapace was used for FPCA.

### Class imbalance and sensitivity analyses

To address class imbalance, we also fit weighted random forest models with class weights inversely proportional to class frequencies, thereby penalizing misclassification of the minority class more strongly. As a demographic sensitivity check, we repeated model fitting in the second-generation FHS participants (Offspring and Omni 1), who are generally older, to assess discrimination within an older cohort.

### Evaluation metrics and uncertainty

Prediction performance was summarized by the area under the receiver operating characteristic curve (AUC), sensitivity, specificity, and their 95% confidence intervals (CIs). The decision threshold for sensitivity/specificity was set by Youden’s index, chosen within training folds and then applied to validation predictions. We also report mean absolute Shapley values across training folds as measures of variable importance. [25]

Plots were created using R packages ggplot2 (version 3.5.1) [26], ggbreak (version 0.1.2) [27], ggpubr (version 0.6.0) [28], and ComplexHeatmap (version 2.15.4) [29].

## 3 Results

### Sample Characteristics

Table 1 presents the demographic and clinical characteristics of the participants considered in this study. The average age was 63.6 years (SD: 13.9), with 1,546 (45%) being male, 785 (23%) having at least one APOE4 allele, and 1,957 (57%) having a college education. Out of the eligible 3,415 FHS participants, 122 (4%) were reviewed to have MCI and 74 (2%) were reviewed to have dementia of any cause at the time of the dCDT. Participants with cognitive impairment (i.e., those diagnosed with MCI or dementia) were typically older, had higher proportions of the APOE4 allele, and were less likely to have a college education compared to the cognitively intact participants. The distribution of Framingham cohorts reflects these age differences: 94% of impaired participants were from the Offspring cohort, whereas 51% of cognitively intact participants were from the Third Generation cohort. Among those with adjudicated impairment, median time from diagnosis to dCDT testing was 1.0 years (Q1: 0, Q3: 4.0). dCDT availability was high across conditions with 3,391 participants (99%) completing the command task and 3,292 (96%) completing the copy task.

**Table 1:** Sample demographic characteristics.

| | **Overall**<br>($N = 3415$) | **Dementia**<br>**or MCI**<br>($N = 196$) | **Cognitively**<br>**Intact**<br>($N = 3219$) |
| --- | --- | --- | --- |
| Age, mean (SD) | 63.6 (13.9) | 82.2 (7.33) | 62.4 (13.4) |
| Male, n (%) | 1546 (45.3) | 80 (40.8) | 1466 (45.5) |
| Education, n (%) |  |  |  |
| Less than high school | 61 (1.8) | 13 (6.6) | 48 (1.5) |
| High school | 545 (16.0) | 63 (32.1) | 482 (15.0) |
| Some college | 852 (24.9) | 63 (32.1) | 789 (24.5) |
| College or more | 1957 (57.3) | 57 (29.1) | 1900 (59.0) |
| Framingham cohort, n (%) |  |  |  |
| Original | 9 (0.3) | 6 (3.1) | 3 (0.1) |
| Offspring | 1440 (42.2) | 184 (93.9) | 1256 (39.0) |
| New Offspring Spouse | 32 (0.9) | 1 (0.5) | 31 (1.0) |
| Third Generation | 1652 (48.4) | 1 (0.5) | 1651 (51.3) |
| Omni 1 | 165 (4.8) | 4 (2.0) | 161 (5.0) |
| Omni 2 | 117 (3.4) | 0 (0) | 117 (3.6) |
| APOE4+, n (%) | 785 (23.0) | 59 (30.1) | 726 (22.6) |
| Command clock, n (%) | 3391 (99.3) | 192 (98.0) | 3199 (99.4) |
| Copy clock, n (%) | 3292 (96.4) | 191 (97.4) | 3101 (96.3) |

### 3.1 Functional Features

Participants with cognitive impairment exhibited more rapidly increasing *G*-functions than cognitively intact participants (Figure 2), indicating greater local clustering (i.e., points more often have a neighbor within a small radius) and less even distribution for participants with cognitive impairment. This pattern was evident in both test conditions; moreover, for each cognitive group, the *G*-function for the command condition increased slightly more rapidly compared to the copy condition.

**Figure 2:**
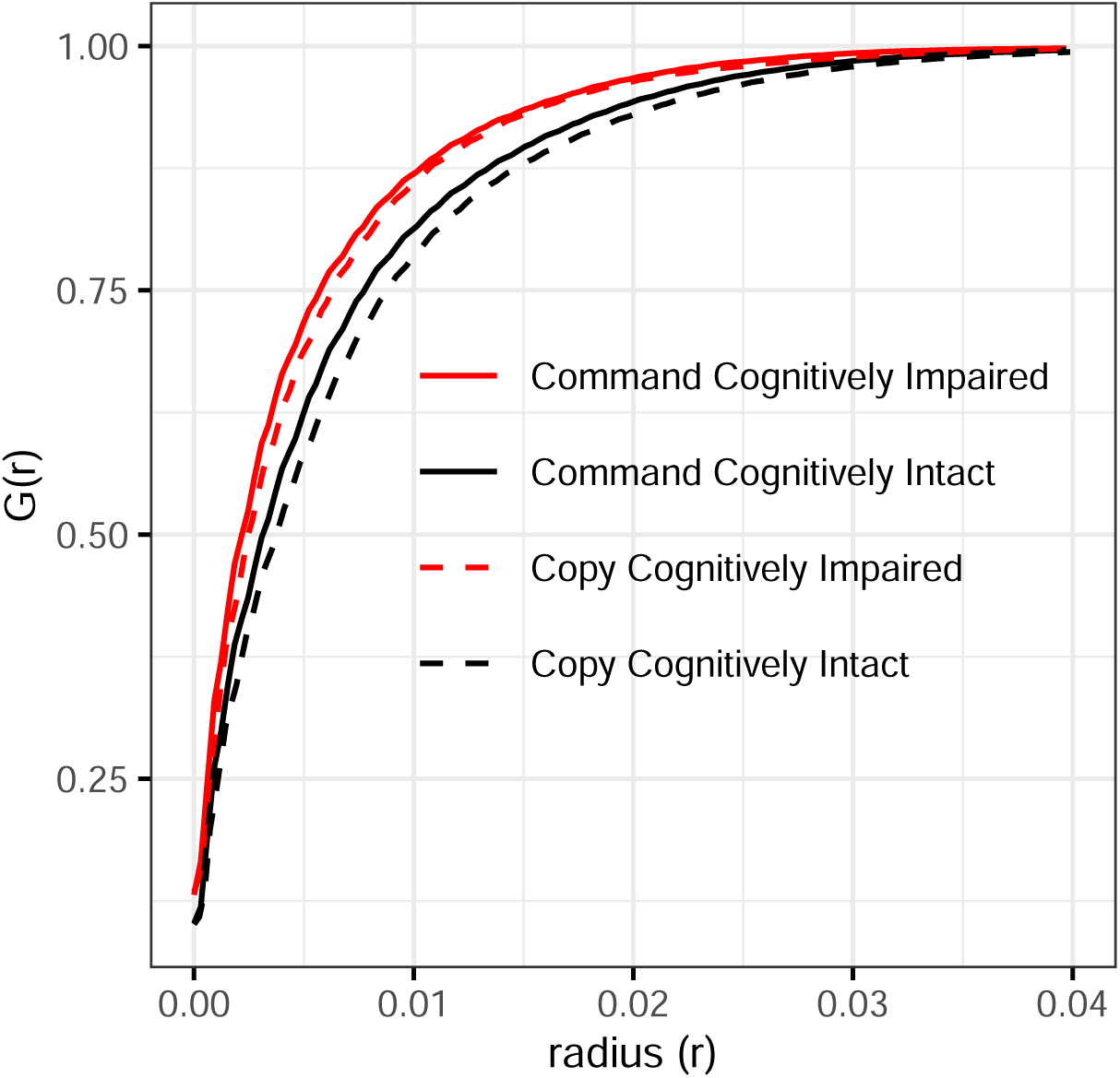
Average G-functions for cognitively intact and cognitively impaired participants in the command and copy test conditions. The G-function is derived from all points recorded by the dCDT to assess spatial distribution. A rapidly increasing function indicates a higher concentration of points in close proximity to each other, reflecting greater clustering of points.

Cognitively impaired participants, on average, exhibited greater variability in the radius function during the command task than intact participants (Figure 3). Because a perfect circle corresponds to a flat radius function, increased variability reflects local deviations from circularity (e.g., asymmetry, waviness, tremors) and imprecision in tracing the clock-face. In the copy task, cognitively impaired participants produced drawings with smaller overall radius values, indicating smaller clock-faces. This size difference is reflected in the average reconstructed clock-faces (Figure 4), where the radius functions were translated into polar coordinates to depict average overall size and shape of the drawings. Clock-faces drawn in the copy condition were generally smaller for both cognitive groups.

**Figure 3:**
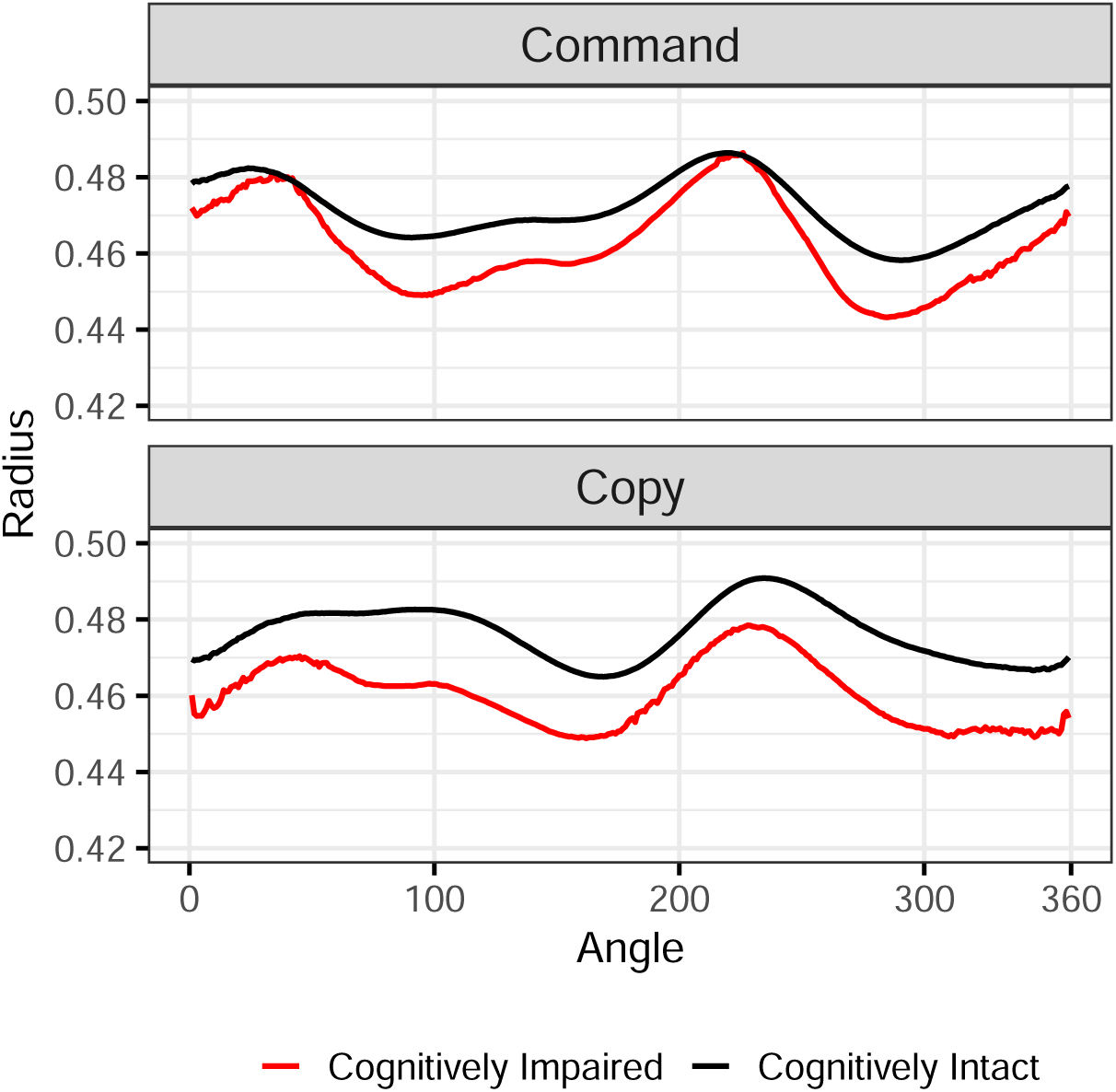
Average radius functions for cognitively intact and cognitively impaired participants in the command and copy test conditions. The radius function is derived from points comprising the clock-face to assess circularity. There is greater variability within the function among cognitively impaired participants indicating greater deviations from circularity.

**Figure 4:**
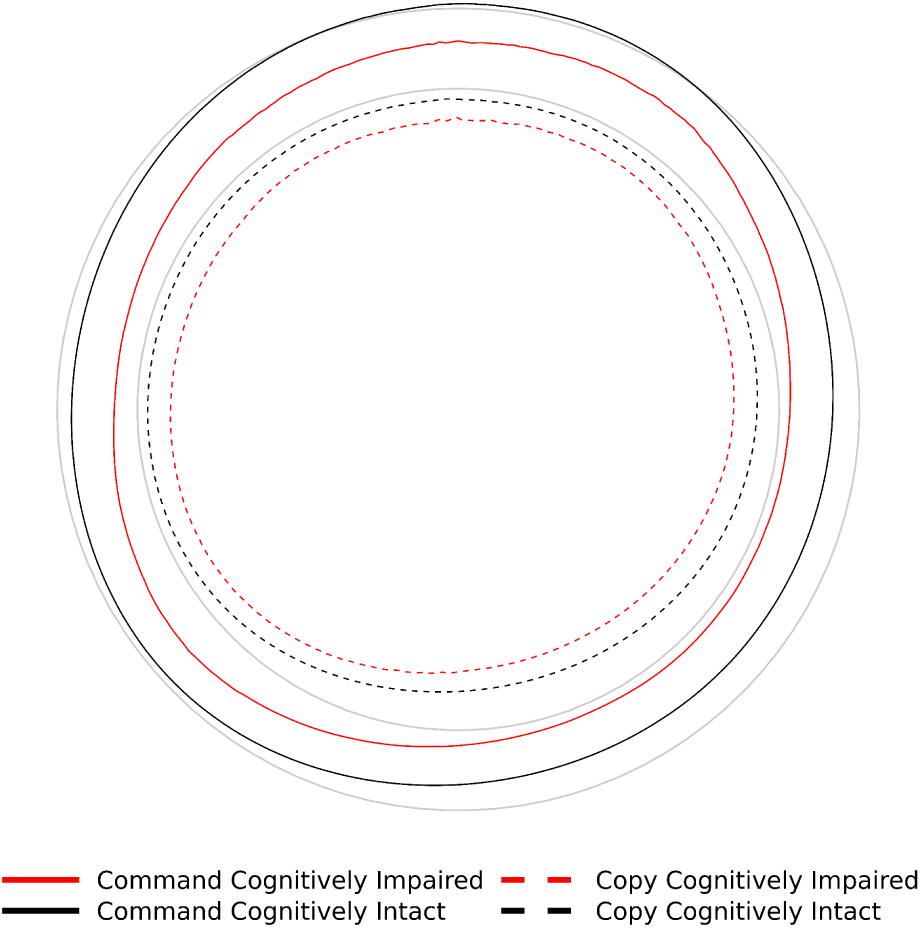
Average clock-faces for participants with and without cognitive impairment after translating the average radius functions into polar coordinates. Note that these average clock-faces are derived by applying the mean scaling factor (see Methods section for details on data processing) from all participant clocks across the two test conditions. Reference lines are in grey.

On average, the pressure density functions were broadly similar between the cognitively intact and impaired participants (Figure 5). The functions were close to zero for values over the central portion of the pressure range (approximately 10–110 on the native scale), with only subtle between-group differences observed in the tails. Relative to intact participants, the impaired group contributed fewer observations at the highest pressures in both conditions and slightly more low-pressure observations in the command condition, suggesting muted peak force during demanding segments of the task. These small tail differences indicate that pressure, when summarized over the full distribution, provides limited separation between groups compared with spatial/morphological signals.

**Figure 5:**
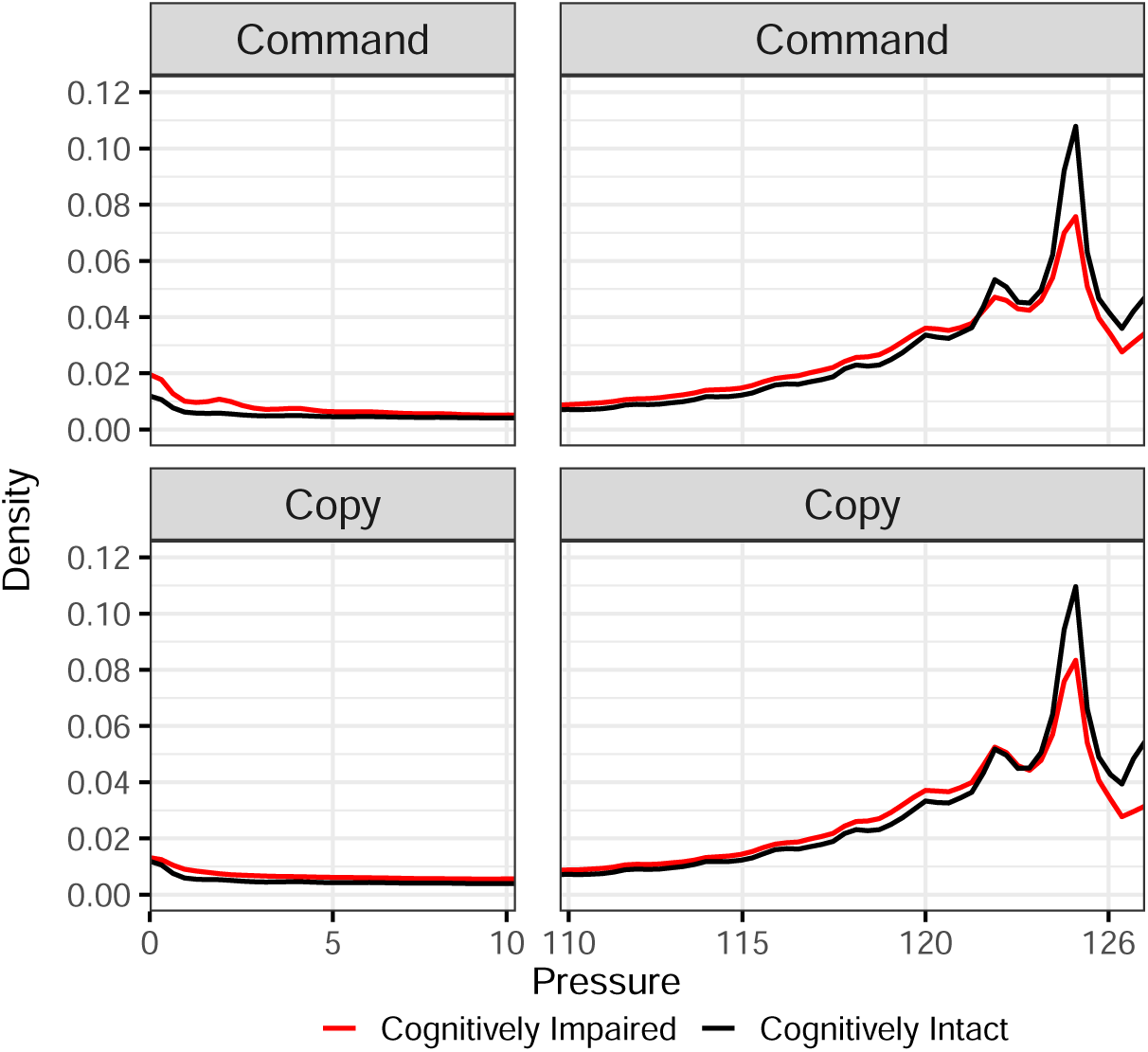
Average pressure density functions for cognitively intact and cognitively impaired participants in the command and copy test conditions. The density function is derived using pressure values from all points recorded by the dCDT to assess variability of pressure applied to the pen for the duration of the test. Note that the plot is divided with a break between pressure values of 10 and 110 to enhance visualization as the primary differences are located in the tails of the distribution.

Overall, these qualitative distinctions support the modeling results (presented next), where functional measures of circularity and spatial proximity provided the clearest group separation.

### 3.2 Cognitively Intact vs. Impaired

Using features from the command task alone (*N* = 3,391), both the *G*-function and radius function demonstrated similar AUC, higher sensitivity, yet reduced specificity to models that incorporated summary measures (Table 2, Figure S1). The *G*-function yielded the highest sensitivity among these models (0.91 [95% CI 0.86–0.94]) but at the cost of lower specificity (0.76 [0.74–0.77]), reflecting its emphasis on detecting clustered spatial behavior characteristic of impairment. The radius-function model balanced performance (sensitivity 0.86 [0.81–0.91], specificity 0.81 [0.80–0.82]) and matched the summary-feature AUC, indicating that circularity deviations capture much of the signal carried by large engineered feature sets. The pressure-density model had slightly lower performance (AUC 0.88) with sensitivity comparable to summary features (0.83) and modest specificity (0.79).

**Table 2:** Performance metrics of random forest models in distinguishing between cognitively normal and cognitively impaired participants.

| Model | N | AUC<br>(95% CI) | Sensitivity<br>(95% CI) | Specificity<br>(95% CI) |
| --- | --- | --- | --- | --- |
| *Command Only* |  |  |  |  |
| Demographics | 3391 | 0.82 (0.79, 0.86) | 0.82 (0.77, 0.87) | 0.78 (0.76, 0.79) |
| Demographics and summary features | 3255 | 0.91 (0.89, 0.93) | 0.83 (0.77, 0.88) | 0.85 (0.84, 0.86) |
| Demographics, time, G-function | 3391 | 0.90 (0.88, 0.92) | 0.91 (0.86, 0.94) | 0.76 (0.74, 0.77) |
| Demographics, time, pressure | 3391 | 0.88 (0.86, 0.90) | 0.83 (0.77, 0.88) | 0.79 (0.78, 0.80) |
| Demographics, time, radius | 3391 | 0.91 (0.89, 0.93) | 0.86 (0.81, 0.91) | 0.81 (0.80, 0.82) |
| Demographics, time, G-function, pressure density, and radius function | 3391 | 0.90 (0.88, 0.92) | 0.89 (0.84, 0.93) | 0.76 (0.75, 0.78) |
| *Copy Only* |  |  |  |  |
| Demographics | 3292 | 0.83 (0.80, 0.86) | 0.81 (0.75, 0.86) | 0.79 (0.78, 0.80) |
| Demographics and summary features | 3225 | 0.90 (0.88, 0.92) | 0.89 (0.84, 0.93) | 0.76 (0.74, 0.77) |
| Demographics, time, G-function | 3292 | 0.90 (0.88, 0.92) | 0.90 (0.85, 0.94) | 0.75 (0.73, 0.77) |
| Demographics, time, pressure | 3292 | 0.88 (0.86, 0.90) | 0.83 (0.77, 0.88) | 0.76 (0.75, 0.78) |
| Demographics, time, radius | 3292 | 0.91 (0.89, 0.93) | 0.85 (0.80, 0.90) | 0.83 (0.81, 0.84) |
| Demographics, time, G-function, pressure density, and radius function | 3292 | 0.89 (0.87, 0.91) | 0.81 (0.76, 0.86) | 0.85 (0.83, 0.86) |
| *Command and Copy* |  |  |  |  |
| Demographics | 3268 | 0.83 (0.80, 0.86) | 0.82 (0.76, 0.87) | 0.79 (0.77, 0.80) |
| Demographics and summary features | 3091 | 0.90 (0.88, 0.92) | 0.88 (0.82, 0.93) | 0.77 (0.75, 0.78) |
| Demographics, time, G-function | 3268 | 0.90 (0.88, 0.92) | 0.91 (0.87, 0.95) | 0.73 (0.72, 0.75) |
| Demographics, time, pressure | 3268 | 0.86 (0.84, 0.89) | 0.76 (0.70, 0.82) | 0.81 (0.79, 0.82) |
| Demographics, time, radius | 3268 | 0.91 (0.89, 0.93) | 0.87 (0.82, 0.92) | 0.80 (0.79, 0.81) |
| Demographics, time, G-function, pressure density, and radius function | 3268 | 0.89 (0.86, 0.91) | 0.79 (0.73, 0.85) | 0.85 (0.83, 0.86) |

Demographics include age, sex, education, Framingham cohort, and APOE-e4 allele presence. Time-based measures include total time to completion, total ink time, and total think time. Models containing G-function, radius function, and/or pressure density also include the image scale factor.

Combining all three functional features did not improve AUC (0.90) and retained the *G*-function’s lower specificity (0.76).

When restricted to the copy task (*N* = 3,292), the *G*-function again matched the summary-feature model on AUC (0.90 vs. 0.90), sensitivity (0.90 vs. 0.89), and specificity (0.75 vs. 0.76). The radius-function model achieved the highest specificity among functions in this setting (0.83 [0.81–0.84]) while maintaining strong discrimination (AUC 0.91), consistent with the descriptive finding that impaired participants draw less circular, smaller faces in copy. Combining all three functional features enhanced the specificity further but with a cost to sensitivity (AUC 0.89; sensitivity 0.81; specificity 0.85). The pressure-density model showed lower performance.

Integrating features from both tasks (*N* = 3,268) preserved these patterns. The radius-function model attained the highest AUC (0.91) with balanced sensitivity/specificity (0.87/0.80), marginally outperforming the summary-feature model (AUC 0.90; 0.88/0.77). The *G*-function improved sensitivity (0.91) whereas pressure density showed lower performance. A model including all three functional features improved specificity (0.85). Supplementary Figure S1 shows receiver operating characteristic (ROC) curves for models with all feature combinations.

Across conditions, functional models based on spatial proximity (*G*) and circularity (radius) consistently matched the discrimination of models using many summary features, with complementary operating profiles. *G*-function yields higher sensitivity (screening), and radius yields higher specificity (fewer false positives). Demographics alone achieved AUCs of 0.82–0.83; adding functional features improved discrimination by ∼0.07–0.09 points. Note that the summary-feature comparator is a literature-aggregated set, not a reproduction of any single model; as such, it provides a strong baseline for summary-feature performance under a common pipeline, making the empirical parity of the *G*- and radius-based models especially notable.

### 3.3 All-cause Dementia vs. MCI

Overall discrimination for differentiating all-cause dementia from MCI was modest across model families, with the best performance observed when using *copy* -task features (Table 3). Using copy-only inputs, the summary features model achieved the highest AUC (0.78; 95% CI 0.70–0.85), outperforming command-only and combined-task counterparts (AUCs 0.62 and 0.71, respectively). Among the functional models in the copy condition, the radius function attained AUC 0.73 (0.65–0.81) with balanced operating characteristics (sensitivity 0.68; specificity 0.74). These patterns suggest that copy-task information is more informative for separating dementia from MCI. Given the smaller sample sizes for this secondary endpoint (command *N* = 192, copy *N* = 191, combined *N* = 187) and the clinical proximity of MCI and dementia, overlapping confidence intervals are expected and likely contribute to the limited gains across models. Plots of the mean functional features stratified by MCI and all-cause dementia are shown in Supplementary Figs. S3-S5.

**Table 3:**
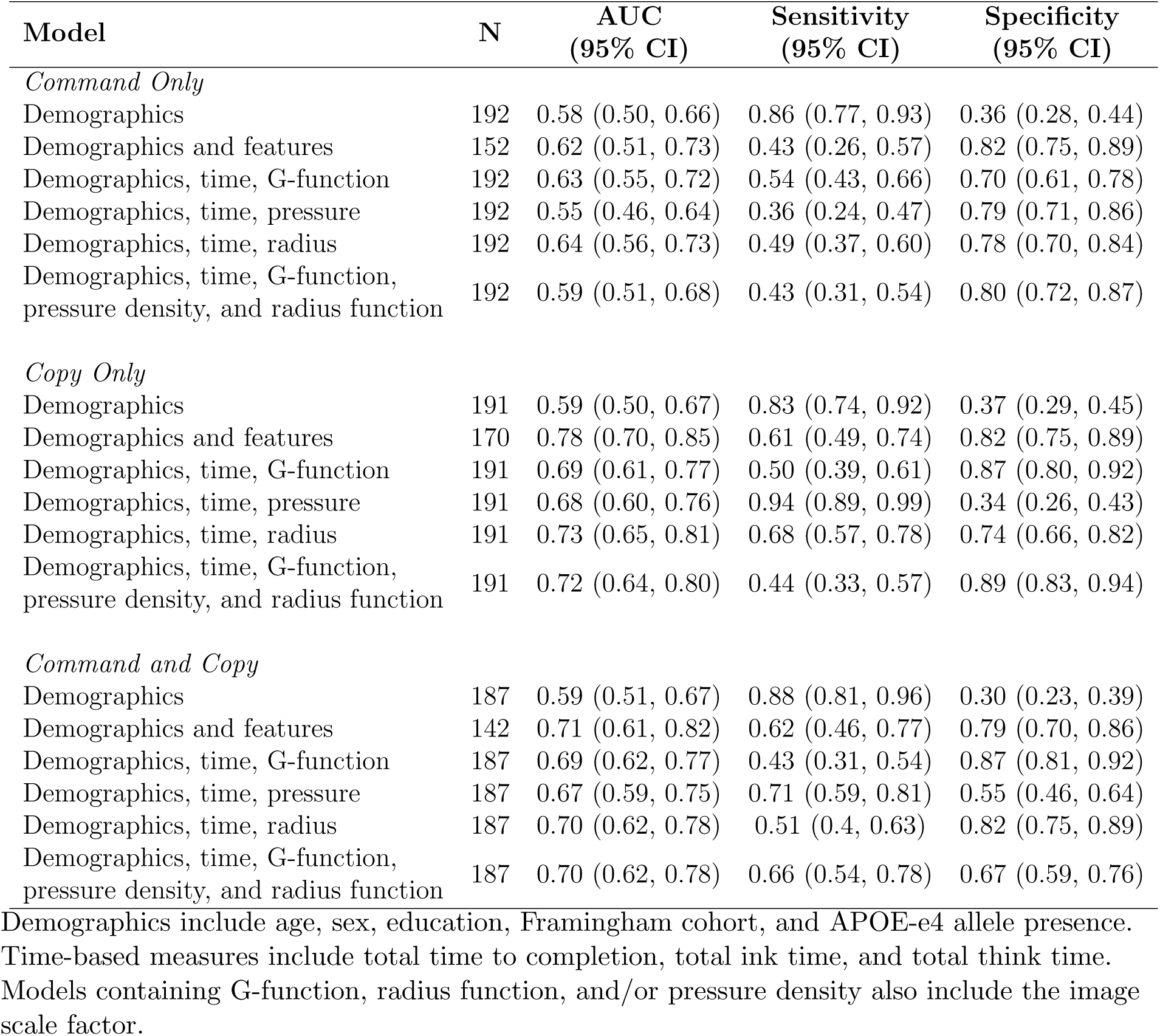
Performance metrics of random forest models in distinguishing between participants with mild cognitive impairment and all-cause dementia.

### 3.4 Sensitivity Analysis

In sensitivity analyses using weighted random forest models to address class imbalance, command, copy or combined dCDT features (summary or functional) provided only minor improvement over the demographics only model (Supplementary Table S2). Weighted random forests primarily shifted the sensitivity–specificity balance rather than substantially changing AUC; sensitivities increased with corresponding decreases in specificity and only minor AUC changes relative to unweighted models.

Restricting to older second-generation participants (Offspring and Omni 1; Supplementary Table S3) expectedly reduced discrimination across models (Supplementary Tables S4–S5). For impaired vs. intact, the summary features model remained strongest or near-strongest across task configurations (AUCs 0.80–0.83), with the radius functional model the most competitive among functional features (AUCs 0.81 command/copy; 0.81 combined) and providing relatively balanced sensitivity/specificity. For the dementia vs. MCI task (Supplementary Table S5), performance was generally modest across all models, with summary features in the copy-only setting again giving the best AUC (0.76). In this older cohort, radius (AUC 0.75 copy-only) provided the most competitive functional alternative.

### 3.5 Variable Importance

Across all primary models, age and FHS cohort consistently ranked as the most influential predictors, followed by time-based measures (completion time, ink time, think time), reflecting known age-related risk and task-duration signals (Supplementary Figure S2). Among functional features, FPC scores from the *G*-function and radius often appeared among the top contributors, aligning with their descriptive separation of groups. However, none of the functional principal component scores for the pressure density function ranked highly in terms of importance. This is consistent with the observed similarities and differences in the averages of these functions between the cognitively impaired and the cognitively intact groups.

## 4 Discussion

The dCDT is a simple, non-invasive, and cost-effective method to detect characteristics of cognitive impairment, providing opportunities for early detection. Our novel features of the dCDT, specifically the *G*-function (spatial proximity) and radius function (circularity) achieved cognitive impairment discrimination comparable to summary-feature models. Relative to demographics alone (AUC: 0.82–0.83), adding either *G*- or radius functions increased AUC by roughly 0.07-0.09 points. These features provide additional insights into drawing behavior relevant to spatial awareness and motor control, beyond what is captured by traditional summary measures.

The *G*-function had similar AUC and sensitivity values to those of the models that used summary features, regardless of test condition. In addition to describing the spatial distribution of points in the test, the *G*-function also provides information into the speed at which the test was completed. Typically, participants who complete the test in a short amount of time have fewer points compared to those who take a longer time. This is reflected in the *G*-function, where larger number of points clustered closely together results in a rapidly increasing function at small radii. Studies have shown that participants with cognitive impairment spend more time with the pen on the paper (i.e., ink time) and require more time to complete the test [8, 11, 12, 14], leading to a greater number of points captured by the pen during the drawing process. This can explain the observation that participants with cognitive impairment in our sample had *G*-functions that increased more rapidly compared to cognitively intact participants. Therefore, the *G*-function holds promise to assess various cognitive domains, including spatial awareness, motor coordination, and processing speed. Models that use *G*(*r*) therefore tend to flag more impaired participants (higher sensitivity), even if this comes with some loss of specificity–an acceptable trade-off for screening, where missed cases are costly.

We observed greater variability in the average radius functions among participants with cognitive impairment in both test conditions (Figure 3). Radius function was as effective as summary features in classifying cognitive status, while typically achieving comparable/higher specificity. The radius function provides a comprehensive, interpretable summary of clock-face circularity, enabling a nuanced understanding of coordination and spatial awareness. A perfectly round clock-face will be reflected in the radius function as a flat line with minimal variability, indicating a symmetrical clock-face with precise and controlled pen strokes. Variability and deviations from a straight line in the radius function arise from non-circular morphology or asymmetry of clock-face, and/or imprecision in the drawing process. Our findings also revealed that participants with cognitive impairment, on average, produced smaller clock-faces in both test conditions compared to those who were cognitively intact (Figure 4). These results align with previous reports that individuals with dementia tend to draw smaller, “avocado-shaped” clocks [17]. Furthermore, we noted that the average size of the clock-faces drawn by all participants was smaller in the copy condition. This is also consistent with previous research showing that cognitively intact adults typically drew clock-faces with smaller areas in the copy condition compared to the command condition [30]. Taken together, variability in the radius function may index differences in visuospatial and executive control, motor coordination, and related cognitive domains beyond what aggregate geometric summaries capture.

The pressure–density functions were broadly similar between cognitively intact and impaired participants across command and copy tasks (Fig. 5). Group differences were confined to the tails, that is, impaired participants contributed fewer observations at the highest pressures in both tasks and slightly more low-pressure observations in the command task. Although models with pressure density had similar AUC values compared to those with summary features, they did not accurately identify individuals with cognitive impairment, as evidenced by the lower sensitivity across all test conditions. This may be due to device-scale constraints on peak pressure, i.e., the pen reaching the maximum pressure limit, preventing recording of higher values that could aid in identifying cognitive impairment.

Age and FHS cohort consistently ranked among the most influential predictors across model specifications, as reflected by their large mean absolute Shapley values (Supplementary Figure S2). This is expected given that cognitive impairment is age-related and older participants tend to be in earlier cohorts. Our analysis explicitly accounted for these demographic factors, along with education and APOE4 genotype to control their confounding effects. Notably, even after this adjustment, functional feature–especially FPC scores from the *G*-function and the radius function–remained informative and improved discrimination over demographics alone, indicating predictive signal beyond well-established demographic risk factors.

Models predicting all-cause dementia from MCI (using either functional or summary features) did not perform as well as combining MCI and dementia into a single cognitively impaired group, as reflected in lower AUC, sensitivity, and specificity. This pattern aligns with the intended use of the CDT as a general cognitive screening tool rather than a diagnostic instrument for separating closely related clinical phenotypes. Even so, the dCDT remains valuable in practice as a simple, non-invasive screening method for identifying individuals who may warrant further evaluation for cognitive impairment.

Addressing class imbalance with weighted random forests did not materially change AUC. And restricting the sample to older participants in sensitivity analyses reduced model performance for all model families, suggesting that variability in age is an key source of discriminative signal. Among the functional features, the radius function remained the most competitive, with balanced operating characteristics in command, copy, and combined settings (Tables S3–S5).

Our feature design and modeling pipeline emphasize interpretability and robustness. Because the *G*-function and pressure density use all recorded pen points and are independent of stroke/component labels, they are robust to clock-symbol segmentation errors. Furthermore, these two features are effectively *test-agnostic*, i.e., they apply not only to the dCDT but also to other digital neuropsychological tests that involve drawing. The radius function depends on the clock-face, but our processing steps allow computation even with overlapping strokes or partially drawn faces. Pre-processing is minimal (scale/translate to the unit square), and each function is summarized by a small set of FPC scores, yielding compact, interpretable predictors that can be visualized at the subject level via loadings and reconstructed curves. To avoid information leakage, we estimate FPCA and select *K* strictly within training folds and project validation curves onto the training-derived eigen- functions; the same stratified cross-validation splits are used for all model families, and thresholds are chosen within folds. We report point estimates with 95% CIs, conduct sensitivity analysis, and quantify importance with Shapley values, supporting a transparent and reproducible analysis. Together, the functional features and the leakage-controlled framework make the approach practical for integration into digital cognitive screening workflows, with modest computational requirements and clear clinical interpretation.

Note that participants not flagged for adjudication were treated as cognitively intact, which may introduce label noise; however, consistent patterns across command/copy tasks and sensitivity analyses suggest robustness in the findings. The cohort is drawn from a single, predominantly White population, which may limit transportability to more diverse settings; external, multi-site validation is warranted. These factors do not alter the central finding that interpretable functional features of spatial proximity and circularity deliver competitive screening performance and remain practical to compute even from incomplete drawings.

Interpretable functional features of spatial proximity (*G*) and circularity (radius) deliver screening performance for cognitive impairment that matches large sets of engineered summary features. The approach requires minimal pre-processing and continues to work when drawings are incomplete, which is often a practical constraint in real-world testing. The *G*-function and pressure-density features are test-agnostic across digital, drawing-based neuropsychological tasks because they use all recorded points and do not depend on component labels; the radius feature is robust to overlaps and partial faces through the convex-hull construction. Across command, copy, and combined inputs, *G* and radius provided complementary operating characteristics suitable for screening, while pressure added little incremental value in this cohort. Together, these results establish functional feature modeling as a principled, practical, and interpretable strategy for digital pen assessments, with clear utility for early identification and triage in cognitive screening.

## Acknowledgments

This work was supported in part by the Framingham Heart Study’s National Heart, Lung, and Blood Institute contract (N01-HC-25195) and by grants from the National Institute of Neurological Disorders and Stroke (R01-NS017950), the National Institute on Aging (U19-AG068753; 1R03AG098551-01) and the Framingham Heart Study Brain Aging Program Pilot Award.

## Author contributions

A.P.: data processing, methodology, formal analysis, interpreting results, writing (original draft), writing (review and editing), visualization. C.K.: data acquisition, data processing, interpreting results, writing (review and editing). Y.T.: interpreting results, writing (review and editing). V.K.: interpreting results, writing (review and editing). K.L: interpreting results, writing (review and editing). S.D.: interpreting results, writing (review and editing). C.L.: interpreting results, writing (review and editing). R.A.: data acquisition, interpreting results, writing (review and editing), funding acquisition. S.M.: conception and design, data processing, methodology, formal analysis, interpreting results, writing (original draft), writing (review and editing), visualization, project supervision and funding acquisition. All authors approve the submitted version of this paper.

## Data availability

Requests to access data from the Framingham Heart Study can be made at https://www.framinghamheartstudy.org/fhs-for-researchers.

## Code availability

The code for this study may be made available on reasonable request from the corresponding author(s).

