## Supplemental Document for "Predicting cognitive impairment using novel functional features of spatial proximity and circularity in the digital clock drawing test"

#### Abstract

This supplement contains additional information on data processing for functional digital clock drawing test features (dCDT), approaches to functional data analysis, definitions of summary dCDT features, and sensitivity analyses restricting participants to second-generation Framingham Heart Study cohorts (Offspring and Omni 1). Figures include mean functional features for the all-cause dementia group and mild cognitive impairment (MCI) group.

### Methods

#### Data processing for radius function

Convex hulls, defined as the smallest convex polygon that encloses a set of points in a plane, were first used to provide an outline of the clock face. This approach eliminates occurrences of "crossing lines" which occur when the end of the participant's path on the clock face intersects with points from the beginning of the path, resulting in visible overlapping lines and thus multiple radii for a given angle. The distance of the points along the convex hull to its centroid were obtained to get the radii. Then the angle of each point relative to the vertical line extending from the centroid was estimated, progressing in a clockwise manner.

The use of convex hulls greatly reduces the number of points along clock face as the resulting polygon is designed to be as small as possible while still containing all of the input points within or on its boundary. Similar to the points along the convex hull, the angle and radius of the remaining points were calculated. In order to use as much of the original data points as possible, the points with non-repeated angles were used along with the convex hull points to comprise the resulting radius function.

The number of points comprising the clock face greatly varied across participants, which resulted in instances where not all integer angle values ranging from 0 to 360 were observed uniformly. Therefore, linear interpolation was used to impute unobserved angle values to improve computational efficiency in statistical analyses.

#### Functional data analysis

When data are observed without error, the coefficients are determined through interpolation, which is accomplished when $K$ equals the number of sampled data points.[1] The coefficients for the basis functions are computed in such a way that yields $x_i(t_j) = y_{ij}$ for all $j$. However, when data are observed with error, the degree of smoothing or noise reduction is controlled by choice of the number of basis functions $K$. The corresponding coefficients are then determined by least squares estimation, which minimizes the quantity $SSE(\mathbf{c}_i) = \sum_{j=1}^{n_i} \left[y_{ij} - \sum_{k=1}^{K} c_{ik}\phi_k(t_{ij})\right]^2 = (\mathbf{y}_i - \mathbf{c}_i^T\boldsymbol{\phi})^T(\mathbf{y}_i - \mathbf{c}_i^T\boldsymbol{\phi})$. The least squares estimate for $\mathbf{c}_i$ is then given by $\hat{\mathbf{c}}_i = (\boldsymbol{\phi}^T\boldsymbol{\phi})^{-1}\boldsymbol{\phi}^T\mathbf{y}_i$.

In the PACE algorithm, following the local linear and quadratic smoothing of the mean and the covariance functions, respectively, the eigenfunctions and eigenvalues of the eigendecomposition are estimated by discretizing the smoothed covariance function, given by $\int \hat{G}(s,t)\hat{\phi}_k(s)ds = \hat{\lambda}_k\hat{\phi}_k(t)$.[2] Consequently, the FPC scores can be approximated by $\hat{\xi}_{ik} = \sum_{j=1}^{N_i}(Y_{ij} - \hat{\mu}(T_{ij}))\hat{\phi}_k(T_{ij} - T_{i,j-1})$.

### Tables and Figures

**Table S1: Descriptions of digital clock drawing test features related to drawing efficiency, information processing, and simple and complex motor skills.**

| Feature | Description | Source |
|---|---|---|
| ***Drawing efficiency*** | | |
| Number of strokes | Total number of strokes used to complete the test | [3, 4] |
| Total time | Total time to complete the test | [3, 5–7] |
| Ink length | Sum of the length of all strokes in the test | [3–5] |
| Strokes per minute | Total number of strokes divided by time | [6] |
| Clock face number of strokes | Total number of strokes used to complete the clock face | [3, 4] |
| Clock face total time | Total time to complete the clock face | [3] |
| Velocity | Drawing ink length divided by total time | [3, 6] |
| ***Information processing*** | | |
| Think time | The amount of time not spent drawing, calculated by subtracting the ink time from total time | [6] |
| Percent think time | Think time divided by total time | [5] |
| Average latency | The average duration of time between each pen stroke | [5] |
| Latency variability | Standard deviation of time between each pen stroke | [5] |
| Post-clock face latency | Delay between finishing the last stroke in the clock face and whatever is drawn next (sec) | [3, 7, 8] |
| Pre-first hand latency | Delay between whatever was drawn before the first clock hand and starting to draw that clock hand (sec) | [3, 7–9] |
| Pre-second hand latency | Delay between whatever was drawn before the second clock hand and starting to draw that clock hand (sec) | [3, 8] |
| Longest latency | The longest time between the end of one stroke and whatever was drawn next | [5] |
| ***Simple and complex motor*** | | |
| Ink time | Total time with the pen on the paper | [3, 6] |
| Percent ink time | Total time with the pen on the paper divided by total time to complete the test | [5] |
| Clock face average speed | Average speed of the pen for all strokes used to complete the clock face | [5] |
| Clock face maximum speed | Maximum speed of the pen when drawing the clock face | [5] |
| Clock face initiation speed | Speed of the pen at the beginning of drawing the clock face | [5] |
| Clock face termination speed | Speed of the pen at the end of drawing the clock face | [5] |
| Average pen pressure | Average pen pressure of all the points in the test | [6] |
| Standard deviation of pressure | Standard deviation of pressure for all points in the test | [3] |

| Feature | Description | Source |
|---|---|---|
| Average pressure/velocity | Ratio of average pressure to average velocity | [3, 6] |
| ***Spatial reasoning*** | | |
| Missing digits | Any digits not appearing on the test | [3] |
| Digit perseveration | Any repeated digits | [3] |
| Missing hands | Missing hour or minute hand | [3] |
| Digit over 12 | Any digits greater than 12 | [3] |
| Hand perseveration | Any repeated minute or hour hand | [3] |
| Hand crossed out | Any hour hand or minute hand that is crossed out | [3] |
| Anchoring | Starting with digits 3,6,9,12 (in any order) before drawing all other digits | [3, 10] |
| Ten-eleven stroke | A line connecting digits 10 and 11 | [3] |
| Clock face area | Area of a circle ($\pi r^2$) using the average radius of horizontal and vertical radii | [3, 7] |
| Hour hand length | Ink length of the hour hand | [3] |
| Hour hand distance from centroid | Distance between the closest point on the hour hand to the centroid of the clock face | [3] |
| Minute hand length | Ink length of the minute hand | [3] |
| Minute hand distance from centroid | Distance between the closest point on the minute hand to the centroid of the clock face | [3] |
| Ratio of minute to hour hand length | Length of minute hand divided by length of hour hand | [3] |
| Average digit height | The average of the heights of all digits drawn on the test | [3] |
| Average digit width | The average of the widths of all digits drawn on the test | [3] |
| Average digit distance from circumference | The average distance from the closest point of each digit to the clock face | [3] |
| Clock face overshoot distance | The minimum distance between the start and end of the clock face divided by the total length of the clock face | [3] |
| Symmetry | Ratio of the length to width of the clock face | [3, 5] |
| Correctly placed hands | Minute and hour hand pointing to the correct digits | [11] |

**Table S2: Performance metrics of weighted random forest models in distinguishing between cognitively intact and impaired participants addressing class imbalance.**

| Model | N | AUC (95% CI) | Sensitivity (95% CI) | Specificity (95% CI) |
|---|---|---|---|---|
| *Command Only* | | | | |
| Demographics | 3391 | 0.85 (0.84, 0.87) | 0.94 (0.90, 0.97) | 0.69 (0.68, 0.71) |
| Demographics and summary features | 3255 | 0.88 (0.85, 0.90) | 0.93 (0.89, 0.97) | 0.72 (0.70, 0.73) |
| Demographics, time, G-function | 3391 | 0.88 (0.86, 0.90) | 0.88 (0.83, 0.92) | 0.76 (0.75, 0.78) |
| Demographics, time, radius function | 3391 | 0.87 (0.85, 0.89) | 0.95 (0.91, 0.97) | 0.69 (0.68, 0.71) |
| Demographics, time, pressure density | 3391 | 0.87 (0.85, 0.89) | 0.85 (0.80, 0.90) | 0.76 (0.75, 0.78) |
| Demographics, time, G-function, radius function, and pressure density | 3391 | 0.86 (0.84, 0.88) | 0.85 (0.80, 0.90) | 0.74 (0.72, 0.75) |
| *Copy Only* | | | | |
| Demographics | 3292 | 0.85 (0.84, 0.87) | 0.91 (0.87, 0.95) | 0.72 (0.70, 0.73) |
| Demographics and summary features | 3225 | 0.87 (0.84, 0.89) | 0.87 (0.82, 0.92) | 0.74 (0.73, 0.76) |
| Demographics, time, G-function | 3292 | 0.87 (0.84, 0.89) | 0.93 (0.88, 0.96) | 0.67 (0.66, 0.69) |
| Demographics, time, radius function | 3292 | 0.86 (0.84, 0.88) | 0.93 (0.89, 0.96) | 0.68 (0.66, 0.70) |
| Demographics, time, pressure density | 3292 | 0.87 (0.85, 0.89) | 0.93 (0.89, 0.96) | 0.66 (0.65, 0.68) |
| Demographics, time, G-function, radius function, and pressure density | 3292 | 0.84 (0.81, 0.86) | 0.92 (0.88, 0.96) | 0.62 (0.61, 0.64) |
| *Command and Copy* | | | | |
| Demographics | 3268 | 0.85 (0.83, 0.87) | 0.94 (0.90, 0.97) | 0.70 (0.68, 0.72) |
| Demographics and summary features | 3091 | 0.85 (0.82, 0.88) | 0.87 (0.82, 0.92) | 0.70 (0.69, 0.72) |
| Demographics, time, G-function | 3268 | 0.87 (0.85, 0.89) | 0.86 (0.80, 0.90) | 0.73 (0.72, 0.75) |
| Demographics, time, radius function | 3268 | 0.85 (0.83, 0.87) | 0.93 (0.89, 0.96) | 0.66 (0.64, 0.68) |
| Demographics, time, pressure density | 3268 | 0.85 (0.82, 0.87) | 0.91 (0.87, 0.95) | 0.66 (0.64, 0.67) |
| Demographics, time, G-function, radius function, and pressure density | 3268 | 0.81 (0.79, 0.84) | 0.90 (0.86, 0.94) | 0.60 (0.59, 0.62) |

Demographics include age, sex, education, Framingham cohort, and APOE4 allele presence. Time-based measures include total time to completion, total ink time, and total think time. Models containing G-function, radius function, and/or pressure density also include the image scale factor.

**Table S3: Sample demographic characteristics of second-generation Framingham participants (Offspring and Omni 1 cohorts).**

| | **Overall** ($N = 1605$) | **Dementia or MCI** ($N = 188$) | **Cognitively Intact** ($N = 1417$) |
|---|---|---|---|
| Age, mean (SD) | 74.2 (8.61) | 81.8 (6.90) | 73.2 (8.32) |
| Male, n (%) | 686 (42.7) | 78 (41.5) | 608 (42.9) |
| Education, n (%) | | | |
| High school or less | 45 (2.8) | 11 (5.9) | 34 (2.4) |
| High school | 319 (19.9) | 60 (31.9) | 259 (18.3) |
| Some college | 446 (27.8) | 63 (33.5) | 383 (27.0) |
| College or more | 795 (49.5) | 54 (28.7) | 741 (52.3) |
| Framingham cohort, n (%) | | | |
| Offspring | 1440 (89.7) | 184 (97.9) | 1256 (88.6) |
| Omni 1 | 165 (10.3) | 4 (2.1) | 161 (11.4) |
| APOE4+, n (%) | 374 (23.3) | 57 (30.3) | 317 (22.4) |
| Command clock, n (%) | 1594 (99.3) | 184 (97.9) | 1410 (99.5) |
| Copy clock, n (%) | 1546 (96.3) | 183 (97.3) | 1363 (96.2) |

**Table S4: Performance metrics of random forest models in distinguishing between cognitively intact vs cognitively impaired participants among second-generation FHS participants (Offspring and Omni 1 cohorts).**

| Model | N | AUC (95% CI) | Sensitivity (95% CI) | Specificity (95% CI) |
|---|---|---|---|---|
| *Command Only* | | | | |
| Demographics | 1594 | 0.74 (0.70, 0.78) | 0.76 (0.69, 0.82) | 0.69 (0.67, 0.71) |
| Demographics and summary features | 1505 | 0.83 (0.80, 0.87) | 0.83 (0.76, 0.88) | 0.71 (0.68, 0.73) |
| Demographics, time, G-function | 1594 | 0.78 (0.75, 0.81) | 0.79 (0.74, 0.85) | 0.64 (0.62, 0.67) |
| Demographics, time, radius | 1594 | 0.81 (0.78, 0.84) | 0.78 (0.72, 0.84) | 0.70 (0.68, 0.72) |
| Demographics, time, pressure | 1594 | 0.75 (0.72, 0.79) | 0.63 (0.57, 0.70) | 0.77 (0.75, 0.79) |
| Demographics, time, G-function, radius function, and pressure density | 1594 | 0.78 (0.75, 0.82) | 0.79 (0.73, 0.85) | 0.64 (0.61, 0.66) |
| *Copy Only* | | | | |
| Demographics | 1546 | 0.73 (0.69, 0.77) | 0.68 (0.62, 0.75) | 0.72 (0.69, 0.74) |
| Demographics and summary features | 1504 | 0.80 (0.76, 0.83) | 0.82 (0.76, 0.88) | 0.64 (0.61, 0.67) |
| Demographics, time, G-function | 1546 | 0.77 (0.73, 0.80) | 0.78 (0.72, 0.84) | 0.65 (0.62, 0.67) |
| Demographics, time, radius | 1546 | 0.81 (0.77, 0.84) | 0.80 (0.74, 0.86) | 0.67 (0.64, 0.69) |
| Demographics, time, pressure | 1546 | 0.79 (0.75, 0.82) | 0.60 (0.52, 0.67) | 0.84 (0.82, 0.86) |
| Demographics, time, G-function, radius function, and pressure density | 1546 | 0.77 (0.74, 0.81) | 0.62 (0.55, 0.69) | 0.78 (0.76, 0.80) |
| *Command and Copy* | | | | |
| Demographics | 1535 | 0.73 (0.69, 0.77) | 0.72 (0.65, 0.78) | 0.69 (0.67, 0.72) |
| Demographics and summary features | 1422 | 0.81 (0.77, 0.85) | 0.78 (0.71, 0.85) | 0.71 (0.69, 0.74) |
| Demographics, time, G-function | 1535 | 0.73 (0.69, 0.77) | 0.73 (0.65, 0.79) | 0.61 (0.58, 0.63) |
| Demographics, time, radius | 1535 | 0.81 (0.78, 0.85) | 0.73 (0.66, 0.79) | 0.76 (0.74, 0.78) |
| Demographics, time, pressure | 1535 | 0.76 (0.72, 0.80) | 0.60 (0.53, 0.66) | 0.82 (0.80, 0.84) |
| Demographics, time, G-function, radius function, and pressure density | 1535 | 0.76 (0.72, 0.80) | 0.59 (0.52, 0.66) | 0.83 (0.80, 0.84) |

Demographics include age, sex, education, Framingham cohort, and APOE-e4 allele presence. Time-based measures include total time to completion, total ink time, and total think time. Models containing G-function, radius function, and/or pressure density also include the image scale factor.

**Table S5: Performance metrics of random forest models in distinguishing between participants with mild cognitive impairment and those with all-cause dementia among second-generation FHS participants (Offspring and Omni 1 cohorts).**

| Model | N | AUC (95% CI) | Sensitivity (95% CI) | Specificity (95% CI) |
|---|---|---|---|---|
| *Command only* | | | | |
| Demographics | 184 | 0.50 (0.42, 0.58) | 0.71 (0.60, 0.83) | 0.41 (0.32, 0.50) |
| Demographics and summary features | 147 | 0.66 (0.56, 0.76) | 0.73 (0.59, 0.86) | 0.60 (0.51, 0.69) |
| Demographics, time, G-function | 184 | 0.60 (0.51, 0.68) | 0.86 (0.78, 0.94) | 0.31 (0.23, 0.4) |
| Demographics, time, radius | 184 | 0.60 (0.52, 0.69) | 0.78 (0.67, 0.87) | 0.41 (0.33, 0.50) |
| Demographics, time, pressure | 184 | 0.55 (0.46, 0.63) | 0.37 (0.24, 0.49) | 0.76 (0.69, 0.83) |
| Demographics, time, G-function, radius function, and pressure density | 192 | 0.59 (0.50, 0.67) | 0.53 (0.41, 0.64) | 0.65 (0.56, 0.73) |
| *Copy only* | | | | |
| Demographics | 183 | 0.54 (0.46, 0.63) | 0.63 (0.52, 0.74) | 0.53 (0.44, 0.62) |
| Demographics and summary features | 164 | 0.76 (0.67, 0.85) | 0.65 (0.52, 0.77) | 0.85 (0.78, 0.91) |
| Demographics, time, G-function | 183 | 0.69 (0.61, 0.78) | 0.57 (0.45, 0.69) | 0.77 (0.69, 0.84) |
| Demographics, time, radius | 183 | 0.75 (0.67, 0.83) | 0.74 (0.63, 0.83) | 0.79 (0.71, 0.86) |
| Demographics, time, pressure | 183 | 0.71 (0.63, 0.79) | 0.68 (0.55, 0.78) | 0.69 (0.60, 0.77) |
| Demographics, time, G-function, radius function, and pressure density | 183 | 0.75 (0.67, 0.82) | 0.68 (0.57, 0.78) | 0.76 (0.69, 0.84) |
| *Command and Copy* | | | | |
| Demographics | 179 | 0.49 (0.40, 0.58) | 0.67 (0.56, 0.79) | 0.41 (0.32, 0.50) |
| Demographics and summary features | 137 | 0.71 (0.61, 0.82) | 0.59 (0.41, 0.76) | 0.78 (0.70, 0.85) |
| Demographics, time, G-function | 179 | 0.70 (0.62, 0.79) | 0.49 (0.36, 0.62) | 0.88 (0.82, 0.94) |
| Demographics, time, radius | 179 | 0.71 (0.62, 0.79) | 0.57 (0.44, 0.70) | 0.80 (0.72, 0.86) |
| Demographics, time, pressure | 179 | 0.68 (0.60, 0.76) | 0.77 (0.66, 0.87) | 0.58 (0.49, 0.67) |
| Demographics, time, G-function, radius function, and pressure density | 179 | 0.70 (0.62, 0.78) | 0.87 (0.79, 0.95) | 0.49 (0.40, 0.58) |

Demographics include age, sex, education, Framingham cohort, and APOE-e4 allele presence. Time-based measures include total time to completion, total ink time, and total think time. Models containing G-function, radius function, and/or pressure density also include the image scale factor.

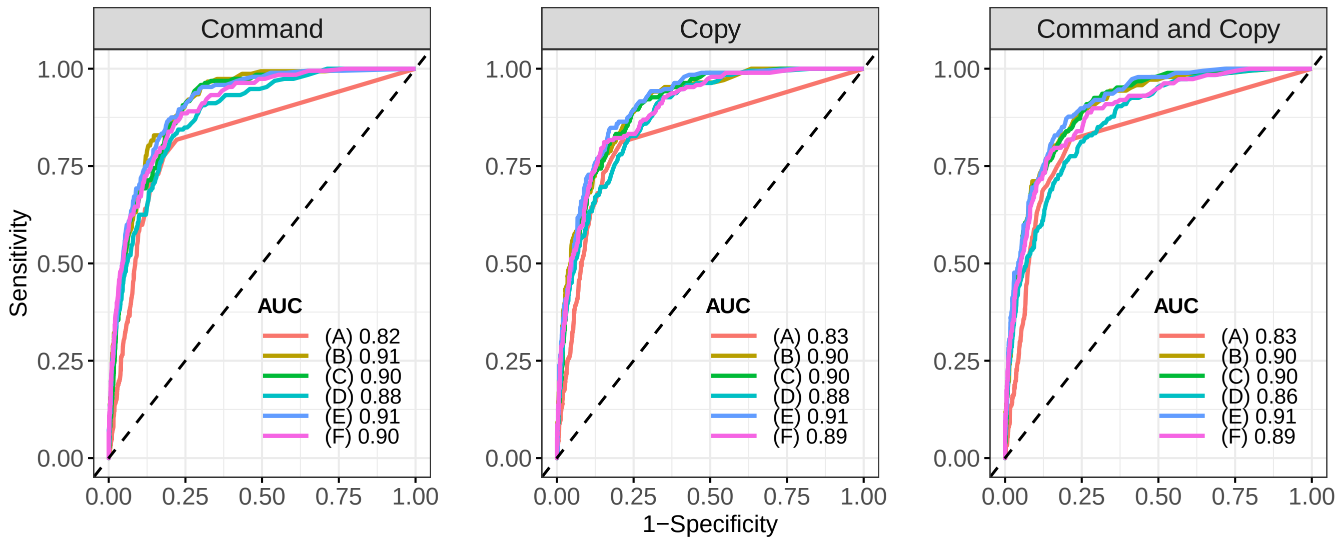


**Fig. S1: Receiver operating characteristic curves for predicting cognitive impairment.** Models contain the following variables: A) demographics and B) summary features; C) G-function; D) pressure density; E) radius function; F) G-function, pressure, and radius functions. Demographics include age, sex, education, Framingham cohort, and APOE4 allele presence. C-F also include image scaling factor, completion time, ink time, and think time.

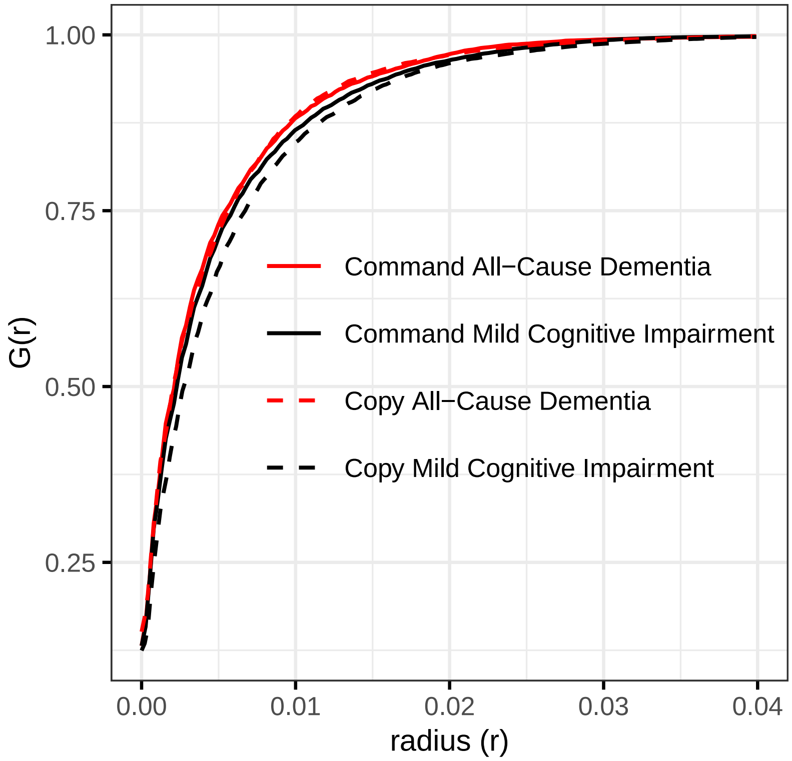


**Fig. S2: Average G-functions for participants with all-cause dementia and mild cognitive impairment in the command and copy test conditions.** The G-function is derived from all points recorded by the dCDT to assess spatial distribution. A rapidly increasing function indicates a higher concentration of points in close proximity to each other, reflecting greater clustering of points.

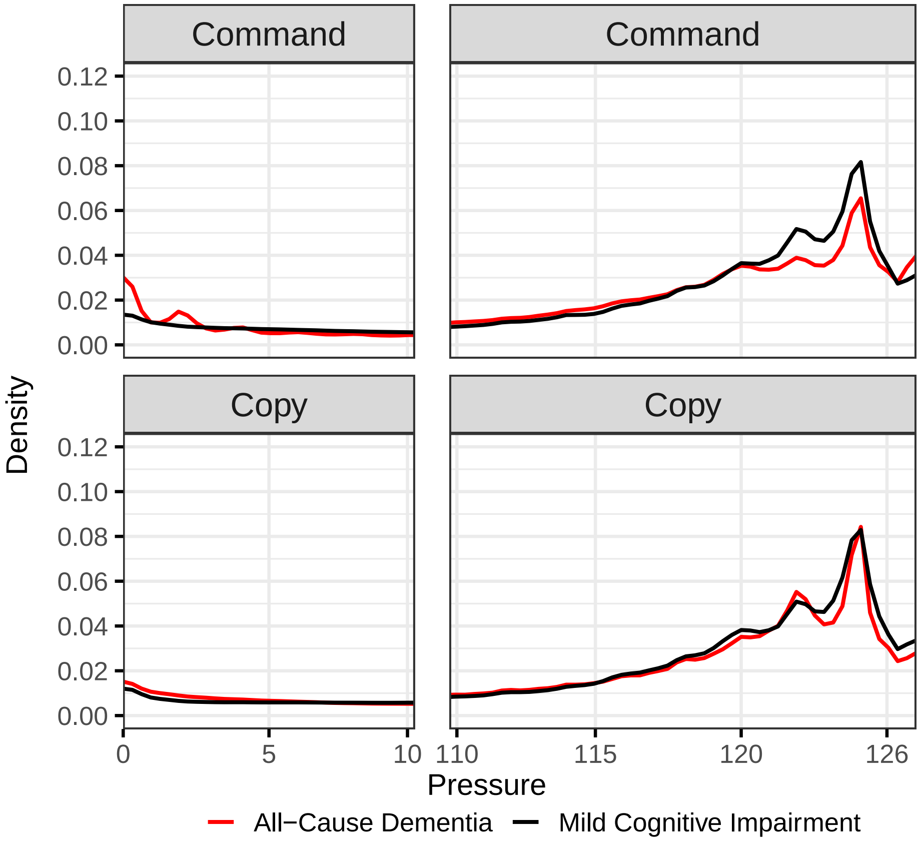


**Fig. S3: Average pressure density functions for participants with all-cause dementia and mild cognitive impairment in the command and copy test conditions.** The density function is derived using pressure values from all points recorded by the dCDT to assess variability of pressure applied to the pen for the duration of the test. Note that the plot is divided with a break between pressure values of 10 and 110 to enhance visualization as the primary differences are located in the tails of the distribution.

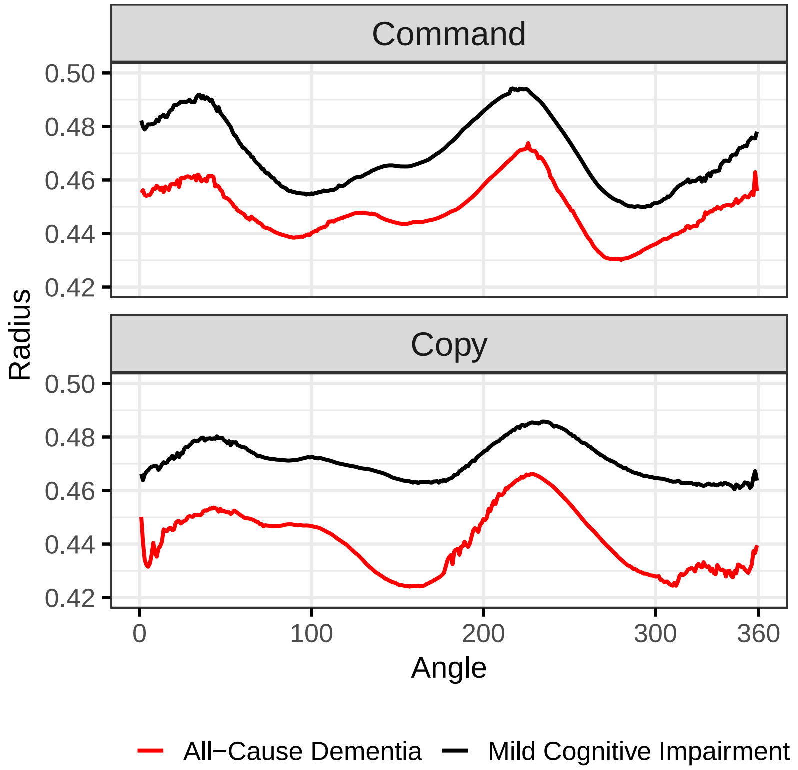


**Fig. S4: Average radius functions for participants with all-cause dementia and mild cognitive impairment in the command and copy test conditions.** The radius function is derived from points comprising the clock face to assess circularity. Participants with dementia of any cause drew smaller clock faces in both test conditions.

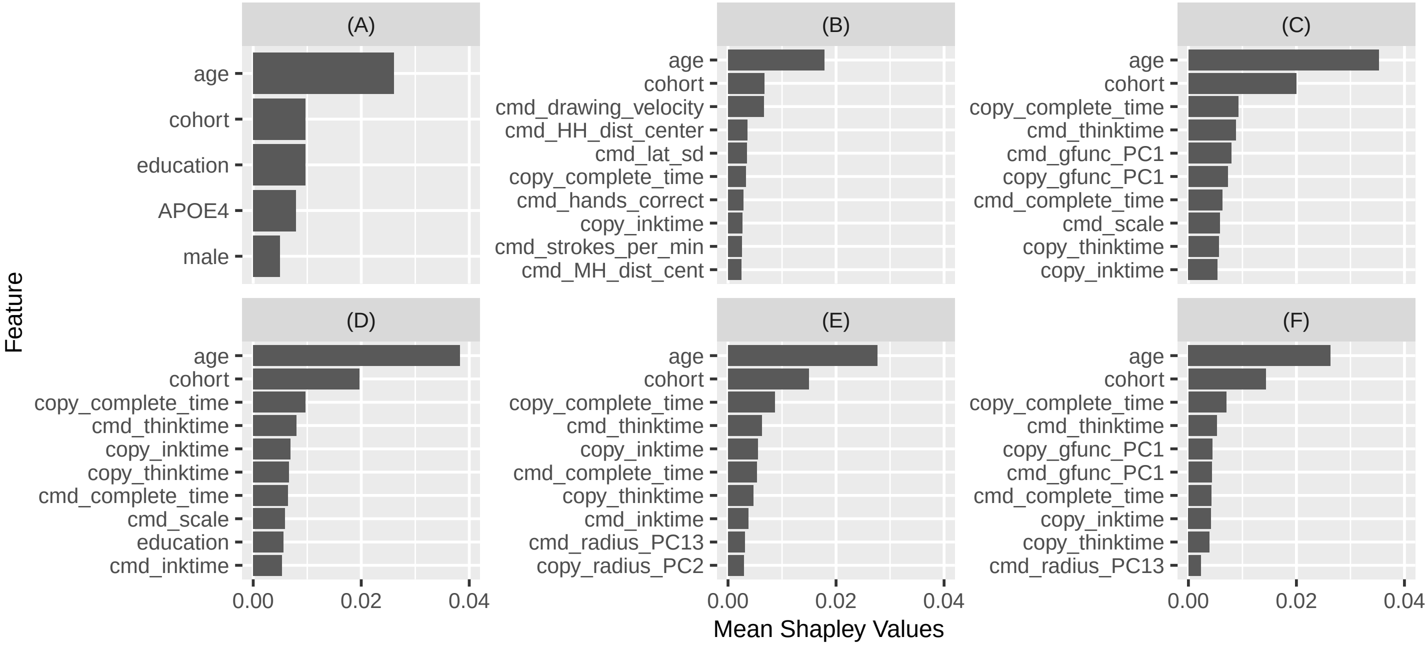


**Fig. S5: Variable importance metrics.** Mean Shapley values across cross-validation folds for features from both command and copy tests. Models contain the following variables: A) demographics and B) summary features; C) G-function; D) pressure density; E) radius function; F) G-function, pressure, and radius functions. Demographics include age, sex, education, Framingham cohort, and APOE-e4 allele presence. C-F also include image scaling factor, completion time, ink time, and think time.
